# Incidence and determinants of fracture risk in 8,348 Mongolian schoolchildren: secondary analysis of data from a randomised controlled trial

**DOI:** 10.1101/2025.10.31.25339221

**Authors:** Barnaby Casson, David A Jolliffe, Sukhbaatar Ariunbuyan, Baigal Delgerekh, Tumenjargal Turmunkh, Uyanga Buyanjargal, Adrian R Martineau, Davaasambuu Ganmaa

## Abstract

**Background:** Data on incidence and determinants of fracture risk among children living in non-Western settings are limited. The predictive value of radial quantitative ultrasound (QUS) for fracture risk in children remains uncertain.

**Methods:** We conducted secondary analyses of data from 8,348 schoolchildren aged 6-13 years living in Ulaanbaatar, Mongolia, who participated in a population-based randomised controlled trial of vitamin D supplementation with 3-year follow-up. Radial speed-of-sound (SOS) was measured at baseline in a subset of 1,456 children. Multivariable Poisson regression was used to estimate fracture incidence, identify determinants of fracture risk, and test whether radial SOS measurements predicted fracture risk.

**Results:** A total of 614 fractures occurred in 521 children over 24,719.3 person-years of follow-up. Overall fracture incidence was 24.9 per 1,000 person-years (95% CI 22.9 to 26.9). Fracture risk was higher in boys vs. girls (adjusted incidence rate ratio [aIRR] 2.10, 95% CI 1.77 to 2.49, P<0.0001) and in older vs. younger children (aIRR 1.18 per additional year of age, 95% CI 1.13 to 1.24, P<0.0001), peaking at age 14 in both boys (87.4 fractures per 1,000 boys per year, 95% CI 63.3 to 117.8) and girls (24.9 fractures per 1,000 girls per year, 95% CI 12.4 to 44.5, respectively). Fracture risk was also higher in children who smoked tobacco vs. those who did not (aIRR 2.20, 95% CI 1.09 to 4.43, P=0.03), while children whose parents were homeowners vs. non-homeowners had lower fracture risk (aIRR 0.83, 95% CI 0.69 to 0.99, P=0.048). Baseline radial SOS did not associate with fracture risk (IRR 0.99; 95% CI 0.99 to 1.00, P=0.31).

**Conclusions:** Fracture incidence among schoolchildren in Mongolia is higher than in Western settings, particularly for boys. Male sex, older age and tobacco smoking were risk factors in this population, while parental home ownership (denoting higher socio-economic status) was protective. Baseline radial SOS did not predict fracture risk in this population.

## INTRODUCTION

An estimated 31 million fractures occurred globally in 2021 among children aged less than 14 years, constituting a substantial source of morbidity, school absence and healthcare expenditure.^1^ Epidemiological studies conducted in Europe and North America have identified increasing age, male sex, pubertal onset, smoking and lower socioeconomic position as risk factors for fractures.^2-9^ Quantitative ultrasound (QUS) has been proposed as a potential low-cost, non-invasive correlate of fracture risk. Cross-sectional studies have reported that radial and tibial speed of sound (SOS) measurements associate with age, pubertal stage and socioeconomic position in adolescents,^10-12^ while case-control studies have reported lower SOS readings in individuals who have experienced a fracture vs. those who have not.^13-15^ However, existing research in this area has largely been conducted in European and North American settings, and data on the predictive value of QUS for incident fracture from low- or middle-income settings, where fracture incidence rates are relatively high,^16^ are lacking.

An opportunity to address this evidence gap arose following completion of a 3-year population-based randomised controlled trial of vitamin D supplementation, which enrolled 8,851 schoolchildren living in Ulaanbaatar, Mongolia, and captured fracture incidence as a secondary outcome.^17^ Details of potential risk factors for fracture were collected at baseline. A subset of 1,456 participants additionally underwent radial SOS measurement at baseline, of whom 1,357 had a repeat measurement at 3-year follow-up. Here, we report results from secondary analyses of trial data, conducted to identify risk factors for incident fractures in this population; to determine whether baseline radial SOS measurements were predictive of subsequent fracture risk; and to evaluate determinants of baseline radial SOS and change in radial SOS over 3-year follow-up.

## METHODS

### Study Design, Setting and Participants

We conducted a parallel two-arm double-blind individually randomised placebo-controlled trial in 18 public schools across Ulaanbaatar, Mongolia, as previously described.^17^ The primary outcome of the trial was acquisition of latent tuberculosis infection, as indicated by conversion of QuantiFERON-TB Gold In-tube (QFT) assay from negative to positive; fracture incidence was a secondary outcome. The current manuscript reports findings from a secondary analysis relating to determinants of fracture risk in the main study population of 8,348 participants, of whom 1,456 underwent radial SOS measurements at baseline. Inclusion criteria for the trial were children aged 6–13 years at time of screening, attending a participating school and the provision of written informed assent from the child and written informed consent from a parent or guardian. Exclusion criteria were a positive QFT assay result, the presence of conditions associated with vitamin D hypersensitivity (e.g. primary hyperparathyroidism or sarcoidosis), having immunocompromised status (e.g. taking immunosuppressive or cytotoxic therapy), the use of vitamin D supplements, clinical signs of rickets, or plans to move out of Ulaanbaatar during the study period. The trial was approved by institutional review boards of the Mongolian Ministry of Health, Mongolian National University, and Harvard T. H. Chan School of Public Health, Boston, USA (ref 14-0513). The trial is registered with clinicaltrials.gov, ref NCT02276755.

### Data collection

Baseline demographic, lifestyle and nutritional data were collected for all 8,348 participants via parent-reported questionnaire, as described elsewhere.^17^ These included: age, sex, ethnicity, outdoor activity, environmental tobacco smoke exposure, active smoking status, family income, level of parental education, home ownership, and type of home and dietary calcium intake. At baseline, 2- and 3-year follow-up, children additionally underwent measurements for height using a portable stadiometer (SECA, Hamburg, Germany), weight using and a digital floor scale (SECA), bioelectric impedance using a body composition analyser (SC-331S, Tanita, Tokyo, Japan), and blood donation for determination of circulating serum 25(OH)D concentrations by VIDAS 25OH vitamin D total enzyme-linked fluorescent assay (bioMérieux, Marcy-l’Étoile, France). At 2- and 3-year follow-up, fracture incidence outcome data were collected by self-reported questionnaires, as previously reported.^18^ Z-scores for height-for-age and BMI-for-age were computed using the 2007 WHO growth reference standards. Non-zero serum 25(OH)D concentrations were standardised using a published method,^19^ utilising a set of 40 reference samples provided by the Vitamin D External Quality Assessment Scheme (DEQAS, http://www.deqas.org/). Values were then adjusted using a sinusoidal model to account for seasonal variation, as previously described.^20^

In 14 participating schools, a subset of 1,465 children was randomly selected to be included in the QUS sub-study and additionally underwent measurement for radial SOS at the distal one-third of the radius using a Sunlight MiniOmni portable bone sonometer (BeamMed, Petah Tikva, Israel), with measurements taken on the non-dominant arm unless the child was injured. Of these, 1,357 had a repeat radial SOS measurement at 3-year follow-up. For each of these participants, change in radial SOS from baseline to follow-up (Δ-SOS) was calculated by subtracting the baseline reading from the follow-up reading.

### Statistical analysis

Data were analysed in Python 3.12.4 with statsmodels 0.14.2 and pandas 2.2.2. Fracture risk was estimated using Poisson models, to allow for repeat fractures and variable follow-up duration, producing incidence rate ratios (IRRs) along with 95% confidence intervals and associated p values. Determinants of baseline radial SOS and Δ-SOS were evaluated using ordinary least squares to fit linear models, producing mean differences along with 95% confidence intervals and associated p values. For all outcomes, univariate factors which were significant at the 10% alpha threshold were carried through to a multivariable model. A multiplication factor of 1000 was applied to radial SOS. Missing outcome data was infrequent: 8,348/8,851 (94.3%) randomised participants contributed fracture outcome data, while 1,357/1,465 (92.6%) of participants who had baseline radial SOS measurements also had follow-up measurements.

## RESULTS

### Participants

Table 1 presents baseline characteristics of the 8,348 trial participants who contributed data to the analysis of determinants of fracture risk, and of the subset of 1,465 participants who also underwent radial SOS measurement. Mean age was 9.4 years (SD 1.6), 49.4% were female and 92.3% were of Khalkh ethnic origin. These and other baseline characteristics in the overall study population were well matched with the subset of participants who contributed SOS data. Total follow-up time was 24,719.3 person-years; average participant follow-up was 2.96 years (SD 0.27).

**Table 1.** Participant characteristics at baseline.

|  |  | Fracture study<br>(n=8,348) | SOS sub-study<br>(n=1,465) |
| --- | --- | --- | --- |
| Mean age, years (s.d.) |  | 9.37 (1.57) | 9.49 (1.56) |
| Sex, *n* (%) | Female | 4,125 (49.4) | 717 (48.9) |
|  | Male | 4,223 (50.6) | 748 (51.1) |
| Ethnicity, *n* (%) | Khalkh | 7,701 (92.3) | 1346 (91.9) |
|  | Other | 647 (7.7) | 119 (8.1) |
| Trial allocation, *n* (%) | Placebo | 4,172 (50.0) | 739 (50.4) |
|  | Intervention | 4,176 (50.0) | 726 (49.6) |
| Height-for-age Z-score (s.d.)<sup>2</sup> |  | -0.28 (0.98) | -0.35 (0.98) |
| BMI-for-age Z-score (s.d.)<sup>2</sup> |  | 0.17 (1.06) | 0.12 (1.04) |
| Body fat, % (s.d.)<sup>2</sup> |  | 17.97 (5.71) | 17.78 (5.68) |
| Radial SOS, (m/s)*1000 (s.d.)<sup>2</sup> |  | --* | 3.64 (0.12) |
| Calcium intake, g/day (s.d.)<sup>2</sup> |  | 0.54 (0.61) | 0.54 (0.62) |
| 25(OH)D concentration, ng/mL (s.d.)<sup>1,2</sup> |  | 11.86 (4.23) | 11.65 (3.97) |
| Outdoor activity, h/day (s.d.)<sup>2</sup> |  | 1.34 (0.84) | 1.19 (0.79) |
| Family income, \$1000s/month (s.d.)<sup>2</sup> | | 0.84 (0.54) | 0.77 (0.44) |
| Parental home ownership, *n* (%) | No | 1,754 (21.0) | 354 (24.2) |
|  | Yes | 6,597 (79.0%) | 1111 (75.8) |
| Type of home, *n* (%) | Centrally heated | 2,026 (24.3) | 228 (15.6) |
|  | Not centrally heated | 3,231 (38.7) | 624 (42.6) |
|  | Ger (yurt) | 3,091 (37.0) | 613 (41.8) |
| Parental education, *n* (%) | University or polytechnic | 3,769 (45.2) | 583 (39.8) |
|  | Secondary school or lower | 4,579 (54.8) | 882 (60.2) |
| Active smoking status, *n* (%)<sup>2</sup> | Non-smoker | 8,302 (99.5) | 1459 (99.6) |
|  | Smoker | 42 (0.5) | 6 (0.4) |
| Environmental tobacco smoke exposure at home, *n* (%)<sup>2</sup> | No | 5,382 (64.5) | 915 (62.5) |
|  | Yes | 2,962 (35.5) | 550 (37.5) |

**Abbreviations:** s.d., standard deviation; BMI, body mass index; SOS, speed of sound 25(OH)D, 25-hydroxyvitamin D.
**Footnotes:** 1, Baseline 25(OH)D values deseasonalised and standardised; to convert to nmol/L, multiply by 2.496. 2, missing data: one missing for height and BMI for fracture study; 36 missing for fat for fracture study, and 5 missing for SOS substudy; 6949 missing for radial SOS for fracture study; 261 missing for calcium intake for fracture study, and 91 missing for SOS substudy; 4 missing for 25(OH)D concentration for fracture study; 4 missing for outdoor activity for fracture study; 1 missing for family income for fracture study and SOS substudy; 4 missing for active smoking and environmental tobacco smoke exposure for fracture study.

### Determinants of fracture risk

A total of 614 fractures occurred in 521 children, of whom 434 experienced one fracture, 81 experienced two fractures and 6 experienced three fractures. Upper limb fractures were most common (436 arising in 374 children), followed by lower limb fractures (138 arising in 134 children) and fractures at other anatomical sites (40 arising in 40 children). Overall fracture incidence rate was 24.9 per 1000 person-years (95% CI 22.9 to 26.9). Determinants of fracture risk are presented in Table 2. Fracture risk was higher with increasing age (adjusted incidence rate ratio [aIRR] 1.18 per additional year of age, 95% CI 1.13 to 1.24, p<0.0001); in males vs. females (aIRR 2.10, 95% CI 1.77 to 2.49, p<0.0001) and in tobacco smokers vs. non-smokers (aIRR 2.20, 95% CI 1.09 to 4.43, p=0.027). Risk of fracture was lower in children whose parents were homeowners vs. non-homeowners (aIRR 0.83, 95% CI 0.69 to 0.99, p=0.048). Figure 2 illustrates that fracture incidence rate (IR) peaked at age 14 both in males (IR 87.4 fractures per 1,000 children per year, 95% CI 63.3 to 117.8) and in females (IR 24.9 fractures per 1,000 children per year, 95% CI 12.4 to 44.5, respectively). Body fat percentage, baseline vitamin D status and level of outdoor activity associated with fracture risk on univariate analysis, but not in multivariable analysis. No other independent variable investigated associated with risk of fracture on univariate analysis, including baseline radial SOS, which was not associated with fracture risk (IRR 0.99; 95% CI 0.99 to 1.00, p=0.31).

**Table 2.** Determinants of fracture incidence.

|  |  | N | Fracture incidence / 1000 ppts/yr | Univariate |  | Multivariate |  |
| --- | --- | --- | --- | --- | --- | --- | --- |
|  |  |  |  | IRR (95% CI) | p | Adjusted IRR (95% CI)<sup>1</sup> | p |
| Age, years |  | 8,348 |  | 1.18 (1.13 to 1.24) | **<0.0001** | 1.17 (1.12 to 1.23) | **<0.0001** |
| Sex | Female | 4,125 | 15.9 | reference |  | reference |  |
|  | Male | 4,223 | 33.7 | 2.10 (1.77 to 2.49) | **<0.0001** | 2.04 (1.71 to 2.43) | **<0.0001** |
| Ethnicity | Khalkh | 7,701 | 25.3 | reference |  |  |  |
|  | Other | 647 | 19.9 | 0.79 (0.57 to 1.10) | 0.17 |  |  |
| Trial allocation | Placebo | 4,172 | 23.9 | reference |  |  |  |
|  | Intervention | 4,176 | 25.9 | 1.09 (0.93 to 1.27) | 0.30 |  |  |
| Height-for-age Z-score |  | 8,347 |  | 0.97 (0.90 to 1.06) | 0.53 |  |  |
| BMI-for-age Z-score |  | 8,347 |  | 0.99 (0.92 to 1.06) | 0.73 |  |  |
| Body fat, % |  | 8,312 |  | 0.98 (0.96 to 0.99) | **0.001** | 1.00 (0.99 to 1.03) | 0.98 |
| Radial SOS, (m/s)*1000 (s.d.) |  | 1,405 |  | 0.99 (0.99 to 1.00) | 0.31 |  |  |
| Calcium intake, g/day |  | 8,090 |  | 0.95 (0.83 to 1.09) | 0.48 |  |  |
| Baseline 25(OH)D concentration, ng/m<sup>2</sup> |  | 8,344 |  | 1.02 (1.00 to 1.04) | **0.032** | 1.01 (1.00 to 1.03) | 0.12 |
| Outdoor activity, h/day<sup>3</sup> |  | 8,344 |  | 1.10 (1.01 to 1.21) | **0.039** | 1.05 (0.96 to 1.16) | 0.27 |
| Family income, \$1000s/month | | 8,347 | | 0.95 (0.81 to 1.11) | 0.54 | | |
| Parental home ownership | No | 1,754 | 28.0 | reference |  | reference |  |
|  | Yes | 6,597 | 24.1 | 0.85 (0.70 to 1.02) | **0.078** | 0.83 (0.69 to 0.99) | **0.048** |
| Type of home | Centrally heated | 2,026 | 23.7 | reference |  |  |  |
|  | Not centrally heated | 3,231 | 24.7 | 1.03 (0.84 to 1.27) | 0.77 |  |  |
|  | Ger (yurt) | 3,091 | 26.0 | 1.08 (0.88 to 1.34) | 0.46 |  |  |
| Parental education | Secondary school or lower | 4,579 | 23.8 | reference |  |  |  |
|  | University or polytechnic | 3,769 | 26.2 | 1.10 (0.94 to 1.29) | 0.24 |  |  |
| Active smoking status | Non-smoker | 8,302 | 24.7 | reference |  | reference |  |
|  | Smoker | 42 | 64.0 | 2.56 (1.28 to 5.15) | **0.008** | 2.20 (1.09 to 4.43) | **0.027** |
| Environmental tobacco smoke exposure at home | No | 5,382 | 24.3 | reference |  |  |  |
|  | Yes | 2,962 | 26.2 | 1.10 (0.93 to 1.29) | 0.28 |  |  |

**Abbreviations:** s.d., standard deviation; BMI, body mass index; SOS, speed of sound; 25(OH)D, 25-hydroxyvitamin D.
**Footnotes:** 1, adjusted for age, sex, body fat %, baseline 25(OH)D concentration, outdoor activity, parental home ownership and active smoking status. 2, baseline 25(OH)D values deseasonalised and standardised; to convert to nmol/L, multiply by 2.496. 3, Outdoor activity was recorded as a categorical variable but analysed as continuous variable, where >2 hours activity per day was converted to 2.5 hours per day.

**Figure 1.**
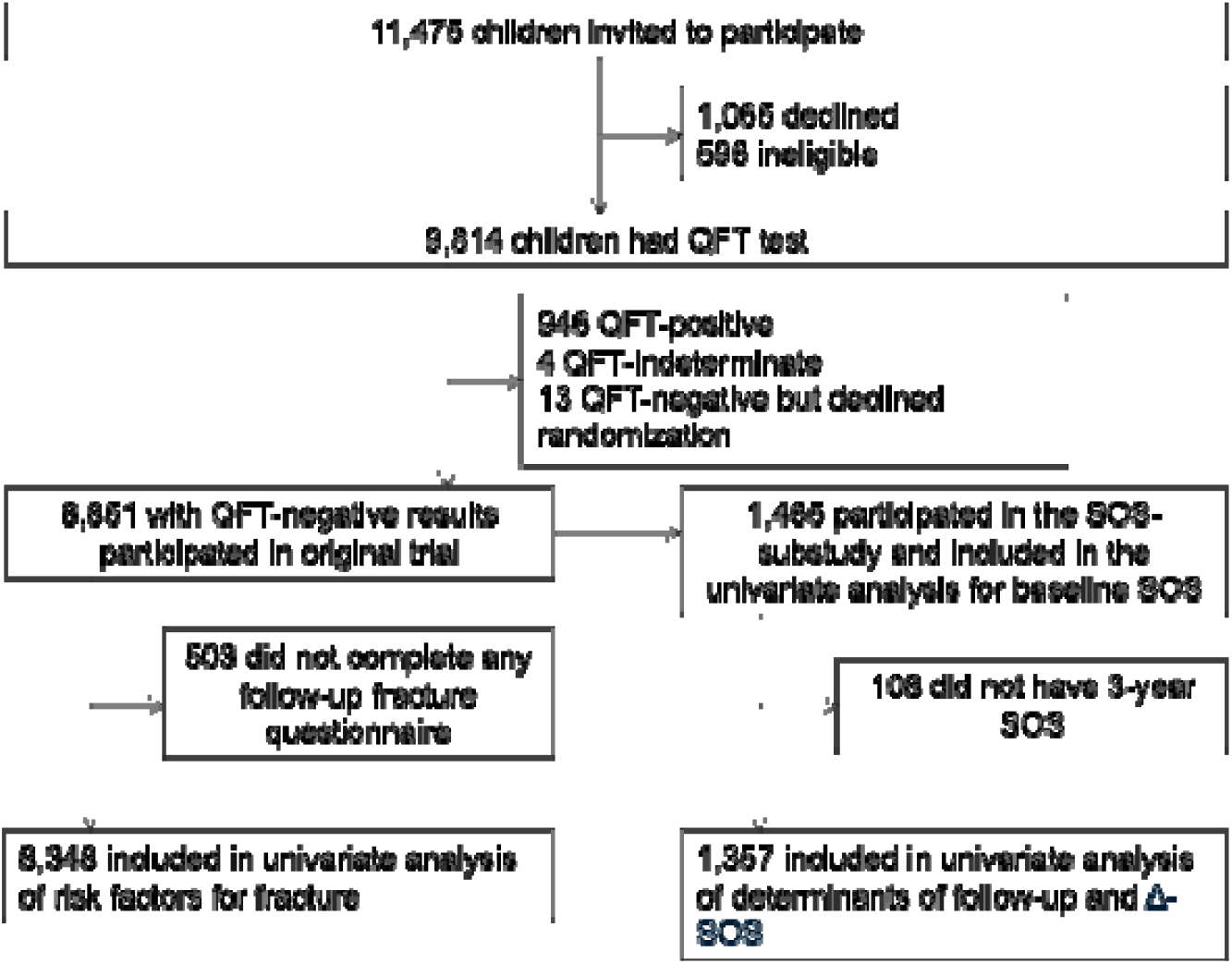
Participant flow. Footnotes: QFT: QuantiFERON-TB Gold in-tube assay; SOS: radial quantitative ultrasound (speed of sound)

**Figure 2.**
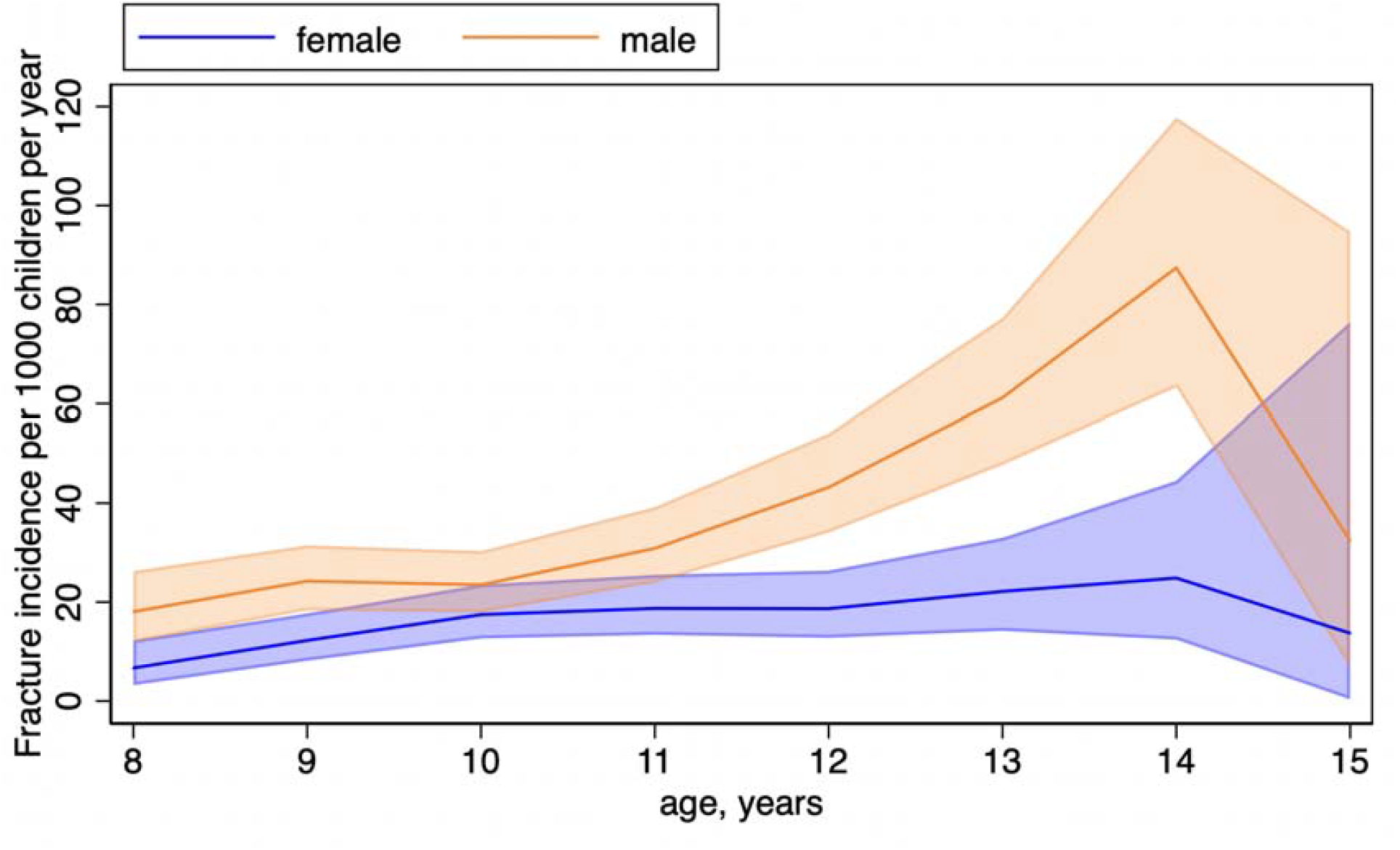
Fracture incidence by age and sex. Lines represent point estimates for fracture incidence rates. Shaded areas represent 95% confidence intervals.

### Determinants of radial SOS

Table 3 presents the determinants of baseline radial SOS. The following factors were independently associated with baseline radial SOS: age (adjusted mean difference [aMD] 15.06 m/s per additional year of age, 95% CI 11.12 to 19.00, P<0.0001) and baseline serum 25(OH)D concentration (aMD 2.32 m/s per additional ng/mL, 95% CI 0.77 to 3.86, P=0.003). Determinants of change in radial SOS over 3-year follow-up (Δ-SOS) are presented in Table 4. Δ-SOS was positively associated with height-for-age Z-score (aMD 15.41m/s per additional Z-score point, 95% CI 7.44 to 23.38, p=.0002) and was lower in males vs. females (aMD −79.82 m/s, 95% CI −94.84 to - 64.80, P<0.0001). Percentage body fat and level of parental education associated with Δ-SOS in univariate analysis but did not remain significant after fitting them into the multivariable model.

**Table 3.** Determinants of baseline radial speed of sound.

|  |  | Mean radial SOS (SD), m/s | Univariate |  |  | Multivariate |  |
| --- | --- | --- | --- | --- | --- | --- | --- |
|  |  |  | n | Mean difference (95% CI) | p | Adjusted mean difference (95% CI) <sup>1</sup> | p |
| Age, years |  |  | 1465 | 14.75 (10.81 to 18.69) | <0.0001 | 15.06 (11.12 to 19.00) | <0.0001 |
| Sex | Female | 3636 (121.0) | 717 | Ref |  |  |  |
|  | Male | 3634 (122.6) | 748 | -2.16 (-14.65 to 10.33) | 0.73 |  |  |
| Ethnicity | Kalkh | 3635 (119.7) | 1346 | Ref |  |  |  |
|  | Other | 3640 (144.2) | 119 | 5.67 (-17.18 to 28.53) | 0.63 |  |  |
| Trial allocation | Placebo | 3632 (120.8) | 739 | Ref |  |  |  |
|  | Intervention | 3638 (122.8) | 726 | 6.56 (-5.92 to 19.05) | 0.30 |  |  |
| Height-for-age Z-score |  |  | 1465 | 0.80 (-5.58 to 7.18) | 0.80 |  |  |
| BMI-for-age Z-score |  |  | 1465 | 1.23 (-4.79 to 7.26) | 0.69 |  |  |
| Body fat, % |  |  | 1460 | 0.56 (-0.54 to 1.66) | 0.32 |  |  |
| Calcium intake, g/day |  |  | 1374 | 1.06 (-9.43 to 11.54) | 0.84 |  |  |
| 25(OH)D concentration <sup>3</sup>, ng/mL |  |  | 1465 | 2.01 (0.44 to 3.58) | **0.012** | 2.32 (0.77 to 3.86) | **0.003** |
| Outdoor activity, h/day |  |  | 1465 | -2.82 (-10.68 to 5.04) | 0.48 |  |  |
| Family income, \$1000s/month | | | 1464 | 5.55 (-8.74 to 19.83) | 0.45 | | |
| Parental home ownership | No | 3626 (117.7) | 354 | Ref |  |  |  |
|  | Yes | 3638 (123.0) | 1111 | 11.84 (-2.73 to 26.42) | 0.11 |  |  |
| Type of home | Centrally heated | 3638 (118.2) | 228 | Ref | 0.99 |  |  |
|  | Not centrally heated | 3640 (119.1) | 624 | 1.94 (-16.55 to 20.42) | 0.84 |  |  |
|  | Ger (yurt) | 3629 (125.7) | 613 | -8.50 (-27.03 to 10.03) | 0.37 |  |  |
| Parental education | University or polytechnic | 3636 (117.3) | 583 | Ref |  |  |  |
|  | Secondary school or lower | 3634 (124.7) | 882 | -1.84 (-14.60 to 10.91) | 0.78 |  |  |
| Active smoking status | Non-smoker | 3635 (121.7) | 1459 | Ref |  |  |  |
|  | Smoker | 3629 (149.8) | 6 | -6.31 (-104.07 to 91.46) | 0.90 |  |  |
| Environmental tobacco smoke exposure at home | No | 3638 (121.1) | 915 | Ref |  |  |  |
|  | Yes | 3630 (122.8) | 550 | -7.72 (-20.60 to 5.17) | 0.24 |  |  |

**Abbreviations:** s.d., standard deviation; BMI, body mass index; SOS, speed of sound 25(OH)D, 25-hydroxyvitamin D.
**Footnotes:** 1, Adjusted for age and 25(OH)D concentration. n=1465. 2, "Other" was chosen as the second category for ethnicity because Kalkha is by far the largest ethnic group in Mongolia. 3, The conversion factor to SI units (nmol/L) is 2.496.

**Table 4.**
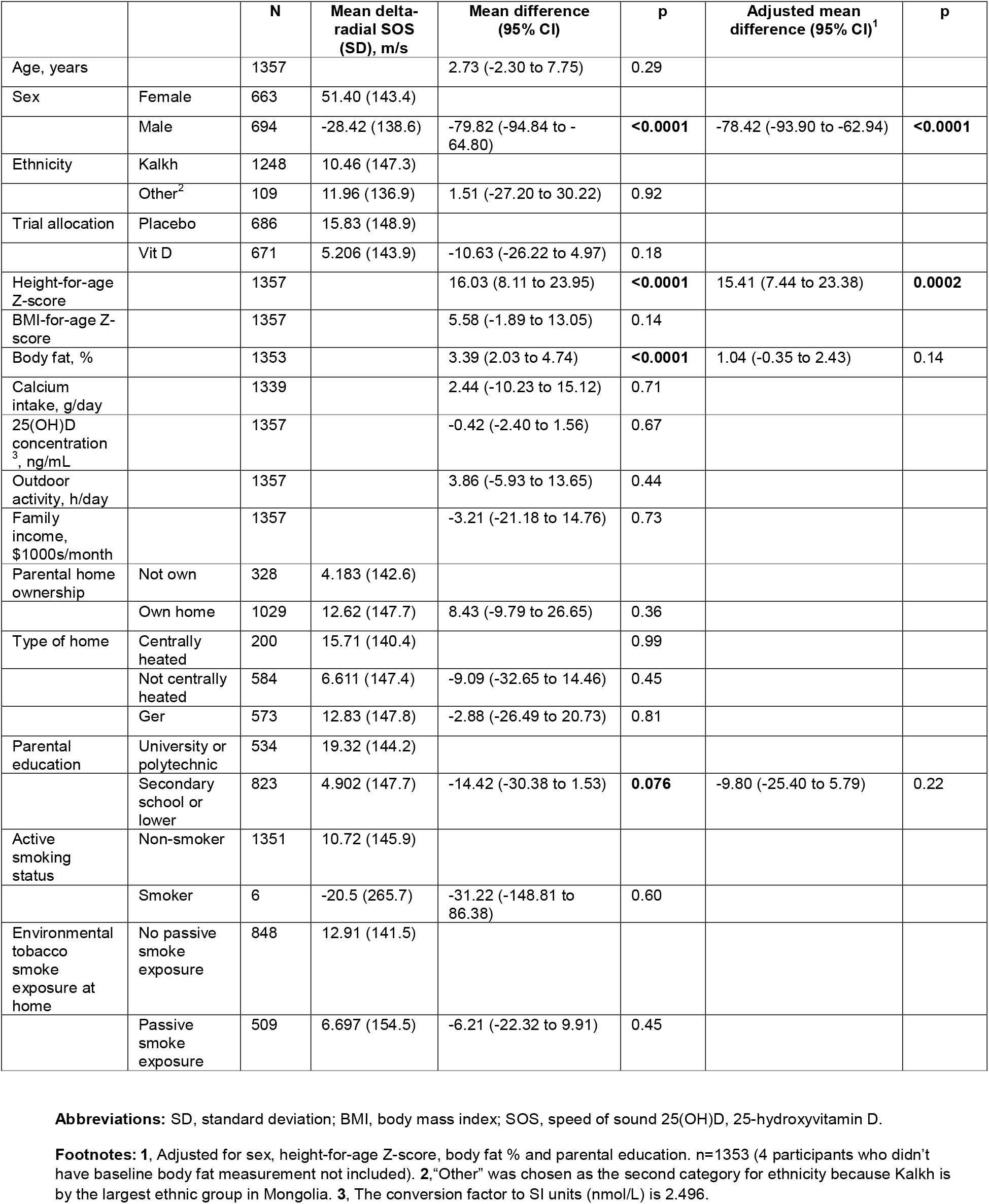
Determinants of delta-radial SOS.

|  |  | **N** | **Mean delta-radial SOS (SD), m/s** | **Mean difference (95% CI)** | **p** | **Adjusted mean difference (95% CI)<sup>1</sup>** | **p** |
| --- | --- | --- | --- | --- | --- | --- | --- |
| Age, years |  | 1357 |  | 2.73 (-2.30 to 7.75) | 0.29 |  |  |
| Sex | Female | 663 | 51.40 (143.4) |  |  |  |  |
|  | Male | 694 | -28.42 (138.6) | -79.82 (-94.84 to -64.80) | **<0.0001** | -78.42 (-93.90 to -62.94) | **<0.0001** |
| Ethnicity | Kalkh | 1248 | 10.46 (147.3) |  |  |  |  |
|  | Other<sup>2</sup> | 109 | 11.96 (136.9) | 1.51 (-27.20 to 30.22) | 0.92 |  |  |
| Trial allocation | Placebo | 686 | 15.83 (148.9) |  |  |  |  |
|  | Vit D | 671 | 5.206 (143.9) | -10.63 (-26.22 to 4.97) | 0.18 |  |  |
| Height-for-age Z-score |  | 1357 |  | 16.03 (8.11 to 23.95) | **<0.0001** | 15.41 (7.44 to 23.38) | **0.0002** |
| BMI-for-age Z-score |  | 1357 |  | 5.58 (-1.89 to 13.05) | 0.14 |  |  |
| Body fat, % |  | 1353 |  | 3.39 (2.03 to 4.74) | **<0.0001** | 1.04 (-0.35 to 2.43) | 0.14 |
| Calcium intake, g/day |  | 1339 |  | 2.44 (-10.23 to 15.12) | 0.71 |  |  |
| 25(OH)D concentration<sup>3</sup>, ng/mL |  | 1357 |  | -0.42 (-2.40 to 1.56) | 0.67 |  |  |
| Outdoor activity, h/day |  | 1357 |  | 3.86 (-5.93 to 13.65) | 0.44 |  |  |
| Family income, \$1000s/month | | 1357 | | -3.21 (-21.18 to 14.76) | 0.73 | | |
| Parental home ownership | Not own | 328 | 4.183 (142.6) |  |  |  |  |
|  | Own home | 1029 | 12.62 (147.7) | 8.43 (-9.79 to 26.65) | 0.36 |  |  |
| Type of home | Centrally heated | 200 | 15.71 (140.4) |  | 0.99 |  |  |
|  | Not centrally heated | 584 | 6.611 (147.4) | -9.09 (-32.65 to 14.46) | 0.45 |  |  |
|  | Ger | 573 | 12.83 (147.8) | -2.88 (-26.49 to 20.73) | 0.81 |  |  |
| Parental education | University or polytechnic | 534 | 19.32 (144.2) |  |  |  |  |
|  | Secondary school or lower | 823 | 4.902 (147.7) | -14.42 (-30.38 to 1.53) | **0.076** | -9.80 (-25.40 to 5.79) | 0.22 |
| Active smoking status | Non-smoker | 1351 | 10.72 (145.9) |  |  |  |  |
|  | Smoker | 6 | -20.5 (265.7) | -31.22 (-148.81 to 86.38) | 0.60 |  |  |
| Environmental tobacco smoke exposure at home | No passive smoke exposure | 848 | 12.91 (141.5) |  |  |  |  |
|  | Passive smoke exposure | 509 | 6.697 (154.5) | -6.21 (-22.32 to 9.91) | 0.45 |  |  |

**Abbreviations:** SD, standard deviation; BMI, body mass index; SOS, speed of sound 25(OH)D, 25-hydroxyvitamin D.
**Footnotes:** 1, Adjusted for sex, height-for-age Z-score, body fat % and parental education. n=1353 (4 participants who didn't have baseline body fat measurement not included). 2, "Other" was chosen as the second category for ethnicity because Kalkh is by the largest ethnic group in Mongolia. 3, The conversion factor to SI units (nmol/L) is 2.496.

## DISCUSSION

In this prospective cohort study of 8,348 Mongolian schoolchildren - which to our knowledge is the largest to investigate the predictive potential of radial quantitative ultrasound for fractures and the first to do so in this setting - we identified (i) a high overall fracture incidence of 24.9 events per 1,000 person-years; (ii) markedly elevated fracture risk in boys, older children and active smokers, with a protective association for parental home-ownership; (iii) no prospective association between baseline QUS measurement and risk of subsequent fractures; (iv) independent associations between higher age and baseline vitamin D status with higher baseline radial SOS; and v) divergent trajectories in radial SOS, showing gains in girls but losses in boys over 3-year follow-up.

Overall fracture incidence in this population was considerably higher than has previously been reported in Europe, ^21-23^ North America,^24^ Africa^25^ and elsewhere in Asia.^26^ Our findings with respect to age and sex as risk factors align with previously published global patterns showing increasing risk in during adolescence, particularly in males.^2,3,7,27^ The protective effect of parental home ownership in the absence of associations for other socioeconomic factors mirrors conflicting findings regarding the influence of socioeconomic deprivation on fracture risk from other settings.^6-8^ Our finding that tobacco smoking is a risk factor for fractures in children is consistent with findings of smaller European and North-American studies that have linked adolescent tobacco use to reduced bone mass and higher fracture risk.^7,8^

Evidence for radial QUS readings as a predictor of fracture risk in children is inconsistent. Early case–control work at phalangeal and tibial sites showed lower SOS in injured participants,^10,11^ whereas recent reviews conclude that paediatric data remain insufficient and heterogeneous, particularly for the radial SOS.^28^ Our null findings extend the literature by providing the first large, prospective test of distal radial SOS in an Asian adolescent population, and reinforce calls to evaluate alternative QUS indices (e.g., broadband ultrasound attenuation, stiffness parameters) or multi-site composites for prediction of fracture risk. The pronounced sex divergence in Δ-SOS over 3-year follow-up (girls +51 m/s vs. boys –28 m/s) likely reflects earlier pubertal onset and oestrogen-mediated cortical consolidation in girls, versus a transient deceleration before the male pubertal growth spurt. The positive associations between baseline vitamin D status and linear growth for baseline radial SOS underscore the sensitivity of QUS to both mineralisation and size-related acoustic pathways.

Our study has several strengths, including a large population size, prospective design, comprehensive characterisation for a range of demographic, anthropometric, biological and biochemical factors and the standardised laboratory calibration of serum 25(OH)D concentrations. There were also some limitations: radial SOS, while a useful proxy for bone stiffness, may not provide a complete picture of bone quality. Bone stiffness indices that also incorporate broadband ultrasound attenuation may reflect bone quality more accurately. Additionally, fractures were self-reported and not radiologically confirmed, introducing possible misclassification.

In conclusion, we report that schoolchildren in Mongolia are at heightened risk of fracture compared with those previously studied in European, North American and other settings, with adolescent boys being at highest risk. Radial quantitative ultrasound readings did not predict fracture risk in this population.

## Data Availability

Anonymized data may be requested from the corresponding authors to be shared subject to terms of IRB approval.

## ACKNOWLEDGEMENTS

This study was supported by an award from the United States National Institutes of Health, ref. 1R01HL122624-01. We thank all the children who participated in the trial, and their parents and guardians. We also thank independent members of the Data Safety Monitoring Board (Prof S.M. Fortune and Dr P.L. Williams, Harvard T.H. Chan School of Public Health; Profs M.F. Holick and C.R. Horsburgh, Boston University; Prof P. Enkhbaatar, University of Texas; and Dr. E. Chadraa, Minnesota State University); independent members of the Trial Steering Committee (Profs W.C. Willett, Edward L. Giovannucci and B.R. Bloom, Harvard T.H. Chan School of Public Health; Dr N. Naranbat, Gyals Medical Laboratory, Ulaanbaatar; and late Dr D. Malchinkhuu, National Center for Maternal and Child Health of Mongolia); board members at the Mongolian Health Initiative (Dr J. Tuyatsetseg, Mongolian University of Science and Technology; Dr J. Amarsanaa, Happy Veritas Laboratory, Ulaanbaatar; Drs P. Erkhembulgan and G. Batbaatar, Mongolian National University of Medical Sciences; Prof M.C. Elliott, Harvard University; and Mr. Ts. Munh-Orgil, Member of the Mongolian Parliament); and the following individuals for advice and helpful discussions: Dr Winthrop Burr (Anadyne Psychotherapy Inc) and Dr. Masae Kawamura at Qiagen USA.

## References

1. Global Burden of Disease Collaborative Network. Global Burden of Disease Study 2021 (GBD 2021). Seattle, United States: Institute for Health Metrics and Evaluation (IHME) Published online 2024.

2. Clark EM. The epidemiology of fractures in otherwise healthy children. Curr Osteoporos Rep 2014; 12(3): 272–8.

3. Rennie L, Court-Brown CM, Mok JY, Beattie TF. The epidemiology of fractures in children. Injury 2007; 38(8): 913–22.

4. Lyons RA, Delahunty AM, Kraus D, et al. Children’s fractures: a population based study. Inj Prev 1999; 5(2): 129–32.

5. Moustaki M, Lariou M, Petridou E. Cross country variation of fractures in the childhood population. Is the origin biological or “accidental”? Inj Prev 2001; 7(1): 77.

6. Stark AD, Bennet GC, Stone DH, Chishti P. Association between childhood fractures and poverty: population based study. BMJ 2002; 324(7335): 457.

7. Jones IE, Williams SM, Dow N, Goulding A. How many children remain fracture-free during growth? a longitudinal study of children and adolescents participating in the Dunedin Multidisciplinary Health and Development Study. Osteoporos Int 2002; 13(12): 990–5.

8. Lyons RA, Delahunty AM, Heaven M, McCabe M, Allen H, Nash P. Incidence of childhood fractures in affluent and deprived areas: population based study. BMJ 2000; 320(7228): 149.

9. Valerio G, Gallè F, Mancusi C, et al. Pattern of fractures across pediatric age groups: analysis of individual and lifestyle factors. BMC Public Health 2010; 10(1): 656.

10. Baroncelli GI, Federico G, Bertelloni S, et al. Assessment of bone quality by quantitative ultrasound of proximal phalanges of the hand and fracture rate in children and adolescents with bone and mineral disorders. Pediatr Res 2003; 54(1): 125–36.

11. Jones G, Boon P. Which bone mass measures discriminate adolescents who have fractured from those who have not? Osteoporos Int 2008; 19(2): 251–5.

12. Schalamon J, Singer G, Schwantzer G, Nietosvaara Y. Quantitative ultrasound assessment in children with fractures. J Bone Miner Res 2004; 19(8): 1276–9.

13. Sosa M, Saavedra P, Gomez-Alonso C, et al. Postmenopausal women with Colles’ fracture have bone mineral density values similar to those of controls when measured with calcaneus quantitative ultrasound. Eur J Intern Med 2005; 16(8): 561–6.

14. Knapp KM, Blake GM, Fogelman I, Doyle DV, Spector TD. Multisite quantitative ultrasound: Colles’ fracture discrimination in postmenopausal women. Osteoporos Int 2002; 13(6): 474–9.

15. Fu Y, Li C, Luo W, Chen Z, Liu Z, Ding Y. Fragility fracture discriminative ability of radius quantitative ultrasound: a systematic review and meta-analysis. Osteoporos Int 2021; 32(1): 23–38.

16. Dong Y, Kang H, Peng R, et al. Global, Regional, and National Burden of Low Bone Mineral Density From 1990 to 2019: Results From the Global Burden of Disease Study 2019. Front Endocrinol (Lausanne) 2022; 13: 870905.

17. Ganmaa D, Uyanga B, Zhou X, et al. Vitamin D Supplements for Prevention of Tuberculosis Infection and Disease. N Engl J Med 2020; 383(4): 359–68.

18. Ganmaa D, Khudyakov P, Buyanjargal U, et al. Vitamin D supplements for fracture prevention in schoolchildren in Mongolia: analysis of secondary outcomes from a multicentre, double-blind, randomised, placebo-controlled trial. The lancet Diabetes & endocrinology 2024; 12(1): 29–38.

19. Sempos CT, Betz JM, Camara JE, et al. General Steps to Standardize the Laboratory Measurement of Serum Total 25-Hydroxyvitamin D. J AOAC Int 2017; 100(5): 1230–3.

20. Ganmaa D, Khudyakov P, Buyanjargal U, et al. Prevalence and Determinants of QuantiFERON-Diagnosed Tuberculosis Infection in 9810 Mongolian Schoolchildren. Clin Infect Dis 2019; 69(5): 813–9.

21. Baig MN. A Review of Epidemiological Distribution of Different Types of Fractures in Paediatric Age. Cureus 2017; 9(8): e1624.

22. Hedstrom EM, Svensson O, Bergstrom U, Michno P. Epidemiology of fractures in children and adolescents. Acta Orthop 2010; 81(1): 148–53.

23. Moon RJ, Harvey NC, Curtis EM, de Vries F, van Staa T, Cooper C. Ethnic and geographic variations in the epidemiology of childhood fractures in the United Kingdom. Bone 2016; 85: 9–14.

24. Naranje SM, Erali RA, Warner WC, Jr., Sawyer JR, Kelly DM. Epidemiology of Pediatric Fractures Presenting to Emergency Departments in the United States. J Pediatr Orthop 2016; 36(4): e45-8.

25. Thandrayen K, Norris SA, Pettifor JM. Fracture rates in urban South African children of different ethnic origins: the Birth to Twenty cohort. Osteoporos Int 2009; 20(1): 47–52.

26. Kamaci S, Yilmaz ET, Goker B, et al. Epidemiology of Pediatric Fractures and Effect of Socioeconomic Status on Fracture Incidence in Turkiye: A Nationwide Analysis of 2 Million Fractures. J Pediatr Orthop 2025; 45(4): e331–e7.

27. Wren TA, Shepherd JA, Kalkwarf HJ, et al. Racial disparity in fracture risk between white and nonwhite children in the United States. J Pediatr 2012; 161(6): 1035–40.

28. Wang KC, Wang KC, Amirabadi A, et al. Evidence-based outcomes on diagnostic accuracy of quantitative ultrasound for assessment of pediatric osteoporosis - a systematic review. Pediatr Radiol 2014; 44(12): 1573–87.

